# Epidemiological Patterns of Melanoma in a Multi-ethnic Cohort in the United Arab Emirates: 2017-2025

**DOI:** 10.64898/2025.12.29.25343157

**Authors:** Jonathan Mokhtar, Nada Alsuwaidi, Nada Hassane, Hind Aljanaahi, Dalia AlDhamin, Tannaz Rahbari, Ghazal Talal Saeed, Zainab Abdulla Al Darwish, Sara Almalik, Jeyaseelan Lakshmanan, Tom Loney, Reem El-Bahtimi

**Affiliations:** Mohammed Bin Rashid University of Medicine and Health Sciences, College of Medicine (MBRU-CoM), Dubai, United Arab Emirates; Department of Dermatology, Kasr Al Ainy School of Medicine, Cairo, Egypt; Department of Dermatology and Aesthetic Center, Rashid Hospital, Dubai Health, Dubai, United Arab Emirates; International DermPath Consult. FZ-LLC, Dubai, United Arab Emirates

**Keywords:** Melanoma, Cancer screening, Public health campaigns, Epidemiology

## Abstract

**Background:** Cutaneous melanoma incidence is rising globally, yet epidemiological data from the high ultraviolet (UV) environment in the United Arab Emirates (UAE), with its diverse expatriate population, remain scarce. This study aims to characterize the epidemiological and histopathological features of cutaneous melanoma in a large, multi-ethnic cohort in the UAE.

**Methods:** This cross-sectional study analyzed histopathologically confirmed cases of cutaneous melanoma diagnosed at a tertiary referral center in the UAE from January 2017 to January 2025. Patient demographics, tumor location, histologic subtype, Clark level, and Breslow thickness were extracted and analyzed. Descriptive statistics, group comparisons, and multivariable logistic regression were performed using IBM SPSS version 29.0 to identify predictors of thick melanoma (Breslow thickness >1.0 mm).

**Results:** A total of 597 patients met the inclusion criteria (50.8% male; mean age 47.4±12.3 years). Individuals of European ancestry constituted 73.4% of cases. Superficial spreading melanoma was the predominant subtype (58.5%), and 46.9% of tumors were thin (≤1.0 mm). Males presented with significantly thicker tumors than females (Breslow thickness of 0.72±1.32 vs. 0.50±0.58 mm; *p <* 0.01) and exhibited distinct anatomical distributions predominant to the back and torso as compared to females with leg and foot predominance. Multivariable analysis identified nodular melanoma (OR 18.40; 95% CI [7.08, 47.86]; *p <* 0.001) and increasing Clark level (OR 18.50; 95% CI [8.44, 40.58]; *p <* 0.001) as strong independent predictors of thick melanoma.

**Conclusion:** Melanoma in the UAE disproportionately affects fair-skinned expatriates and frequently presents with sex-specific clinical patterns. These findings highlight the need for targeted public awareness initiatives to reduce melanoma morbidity and mortality in the region.

## 1. Introduction

Melanoma, a malignant neoplasm of melanocytes, represents a significant and escalating global health challenge. Despite many cases being preventable, it remains the most lethal form of skin cancer worldwide[1]. The global burden of melanoma is substantial, with an estimated 325,000 new cases and 57,000 deaths reported in 2020 alone, with recent GLOBOCAN estimates indicating that in 2025, approximately 107,960 cases of cutaneous melanoma were diagnosed, with 8430 deaths occurring worldwide[2]. Projections indicate a concerning trajectory, with the number of new cases expected to rise by approximately 50% to 510,000 and deaths to increase by 68% to 96,000 by 2040 if current trends persist[2]. The incidence of melanoma exhibits substantial geographic variation, with the highest rates observed in sun-exposed populations in Australia and New Zealand, followed by North America and Europe[2,3]. Conversely, melanoma has historically been considered rare in most African and Asian countries, including the Middle East, with incidence rates often below 1 per 100,000 person-years[2].

However, this traditional view of melanoma as a low-incidence cancer in the Middle East is being challenged by emerging data[4,5]. While research output from the region remains limited, studies suggest a growing number of cases, highlighting a potential shift in the region’s epidemiological landscape[4]. In the United Arab Emirates (UAE), a nation characterized by its multicultural expatriate population, year-round sun exposure, and rapid urbanization, skin cancer now ranks as the fourth most common malignancy[5]. This rising incidence occurs within a context of significant barriers to early detection and prevention.

The confluence of a rising cancer burden and low public awareness creates a critical public health issue. The unique demographic profile of the UAE, with its diverse mix of skin phenotypes and cultural attitudes towards sun exposure, presents a complex scenario for which there is a paucity of specific data. Understanding the distinct epidemiological patterns, risk factors, and clinical presentations of melanoma in this population is essential for developing effective, targeted public health interventions, awareness campaigns, and clinical screening strategies. Therefore, this study was conducted to investigate the epidemiological pattern of melanoma in the UAE, aiming to address existing knowledge gaps and provide a robust evidence base to inform future healthcare policy and practice in the region.

## 2. Materials and Methods

### 2.1 Study design and Setting

This study was conducted as a retrospective cross-sectional analysis, adhering to the Strengthening the Reporting of Observational Studies in Epidemiology (STROBE) guidelines **(Supplementary File 1)**[6]. We retrospectively identified all patients diagnosed with histopathologically confirmed melanoma during the eight-year period from January 1, 2017, to January 1, 2025 (the senior author accessed the data, and de-identified records were available only to the authors of the paper). The study was conducted at a single, high-volume tertiary referral center in the UAE (International DermPath Consult FZ-LLC), a setting characterized by a highly diverse, multinational patient population. The study conformed to the ethical principles of the Declaration of Helsinki, and IRB approval was granted from the Dubai Scientific Research Ethics Committee (DSREC) (Reference number: DSREC-11/2024_41).

### 2.2 Study Population and Participant Selection

The population included all patients who received a new, primary diagnosis of cutaneous melanoma, confirmed by histopathological examination, at our institution during the defined study period. Patients with both invasive melanoma and melanoma in situ were eligible for inclusion to ensure a complete epidemiological profile. Case identification was performed by the senior author, who systematically searched the institution’s electronic pathology database using specified diagnostic codes corresponding to all subtypes of cutaneous melanoma. To ensure the integrity of the cohort, we applied specific exclusion criteria. Patients were excluded if they were diagnosed with non-melanoma skin cancer, presented with metastatic melanoma from an unknown primary site, or had medical records that were incomplete or inaccessible, thereby precluding the reliable extraction of key demographic and clinicopathological variables.

### 2.3 Diagnostic Procedures and Histopathological Assessment

Initial clinical assessment of suspicious pigmented lesions in the UAE was performed by board-certified dermatologists using the ABCDE criteria (Asymmetry, Border irregularity, Color variegation, Diameter >6mm, Evolving)[7]. Patients with lesions meeting these criteria underwent a full-thickness excisional biopsy with a 2-3mm clinical margin, which is the standard of care for definitive diagnosis. Upon receipt in the pathology laboratory, specimens were fixed in 10% neutral buffered formalin, serially sectioned, and entirely submitted for processing. The tissue was then embedded in paraffin, and 4-5 µm-thick sections were cut and stained with hematoxylin and eosin. For diagnostically challenging cases, immunohistochemical staining was performed using a panel of melanocytic markers, including S-100, Melan-A, and HMB-45, to confirm the diagnosis and differentiate melanoma from other pigmented lesions. Prepared slides were subsequently reviewed and diagnosed by the senior author.

### 2.4 Data Collection and Variable Definitions

A standardized, de-identified data collection instrument was developed in Microsoft Excel by the senior author to safeguard patient privacy and data integrity. All relevant variables were systematically extracted from the patients’ electronic health records (EHR) and corresponding definitive pathology reports. Data extraction was performed independently by two authors to ensure accuracy and minimize transcription errors, with any discrepancies adjudicated by the senior author. Demographic characters were recorded as follows: age at diagnoses, captured as a continuous variable (in years), sex was documented as male or female, and self-reported ethnicity was classified into six geographically based categories according to World Health Organization (WHO) designations: European, Australasian, Middle Eastern and North African (MENA), African, South and Southeast Asian, and South American.

All tumor characteristics were based on the final official histopathological report. The histological subtype was classified according to the 2018 WHO classification of skin tumors, including superficial spreading melanoma (SSM), nodular melanoma (NM), lentigo maligna melanoma (LMM), melanoma in situ, and a category for other rare or unclassifiable subtypes (including acral lentiginous melanoma [ALM], desmoplastic melanoma, and spitzoid melanoma)[8]. The primary prognostic indicator, Breslow thickness, was recorded in millimeters (mm) and represents the maximal vertical tumor thickness measured from the top of the granular layer of the epidermis to the deepest point of tumor invasion. For analytical purposes, Breslow thickness was treated as both a continuous variable and was categorized into four clinically relevant groups based on the American Joint Committee on Cancer (AJCC) staging system: ≤1.0 mm (thin), 1.01–2.0 mm, 2.01–4.0 mm, and >4.0 mm (very thick)[9]. The Clark level of invasion, representing the anatomical plane of invasion through the layers of the skin, was recorded as levels I through V[9]. Finally, the tumor’s primary anatomical location was documented and grouped into four major body sites: head and neck, back and torso, arm and hand, and leg and foot.

### 2.5 Bias

Potential biases were addressed through several strategies. Selection bias was minimized by including data from diverse healthcare facilities across several Emirates within the UAE, representing a wide range of demographic groups. To ensure comparability across patient groups, all histopathological analyses were conducted centrally at the International DermPath Consult, FZ-LLC, Dubai, UAE, using standardized protocols, thereby reducing inter-observer variability. Observer bias was further reduced by involving an independent epidemiologist and biostatistician (T.L. and J.L.), who performed the statistical analysis using the de-identified dataset linked only by unique identification numbers, without access to any direct patient identifiers.

### 2.6 Statistical Analysis

All statistical analyses were conducted using SPSS Statistics for Windows, Version 28.0 (IBM Corp., Armonk, NY). The cohort’s characteristics were summarized using descriptive statistics. Continuous variables that were normally distributed, such as age and Breslow depth, were reported as mean ± standard deviation (SD). Categorical variables, including sex, ethnicity, histological subtype, and anatomical location, were reported as frequencies and percentages (%).

To evaluate differences in clinical presentation between sexes, group comparisons were performed. An independent-samples t-test was used to compare the means of normally distributed continuous variables (age and Breslow depth) between males and females. For categorical variables, the chi-square (χ²) test was employed; in cases where the expected cell count was less than five, Fisher’s exact test was used to ensure statistical validity. A two-sided p-value <0.05 was considered statistically significant for all analyses.

To identify independent factors associated with more advanced disease at presentation, a multivariable logistic regression analysis was conducted. The primary outcome for this model was the presence of a thick melanoma, defined as Breslow depth >1.0 mm. The dependent variable was therefore dichotomized into thin (≤1.0 mm) and thick (>1.0 mm) tumors. Covariates selected for inclusion in the model were based on clinical relevance and included age group, sex, primary anatomical body site, histological subtype, and Clark level of invasion. Clark’s level was treated as a continuous variable in the model to prevent convergence issues arising from categorization. The results of the regression analysis were presented as odds ratios (ORs) with their corresponding 95% confidence intervals (CIs). The goodness-of-fit of the final logistic regression model was formally assessed using the Hosmer-Lemeshow test.

## 3. Results

### 3.1 Participants

Over the eight-year study period, 597 patients met the inclusion criteria and were included in the final analysis **(Figure 1)**. Reasons for exclusion of other participants included the following: metastatic disease, simple nevi, and other diagnoses not related to melanoma. The cohort was almost evenly split by sex, comprising 303 males (50.8%) and 294 females (49.2%). The baseline demographic and clinical characteristics of the study population are summarized in **Table 1**.

**Figure 1.**
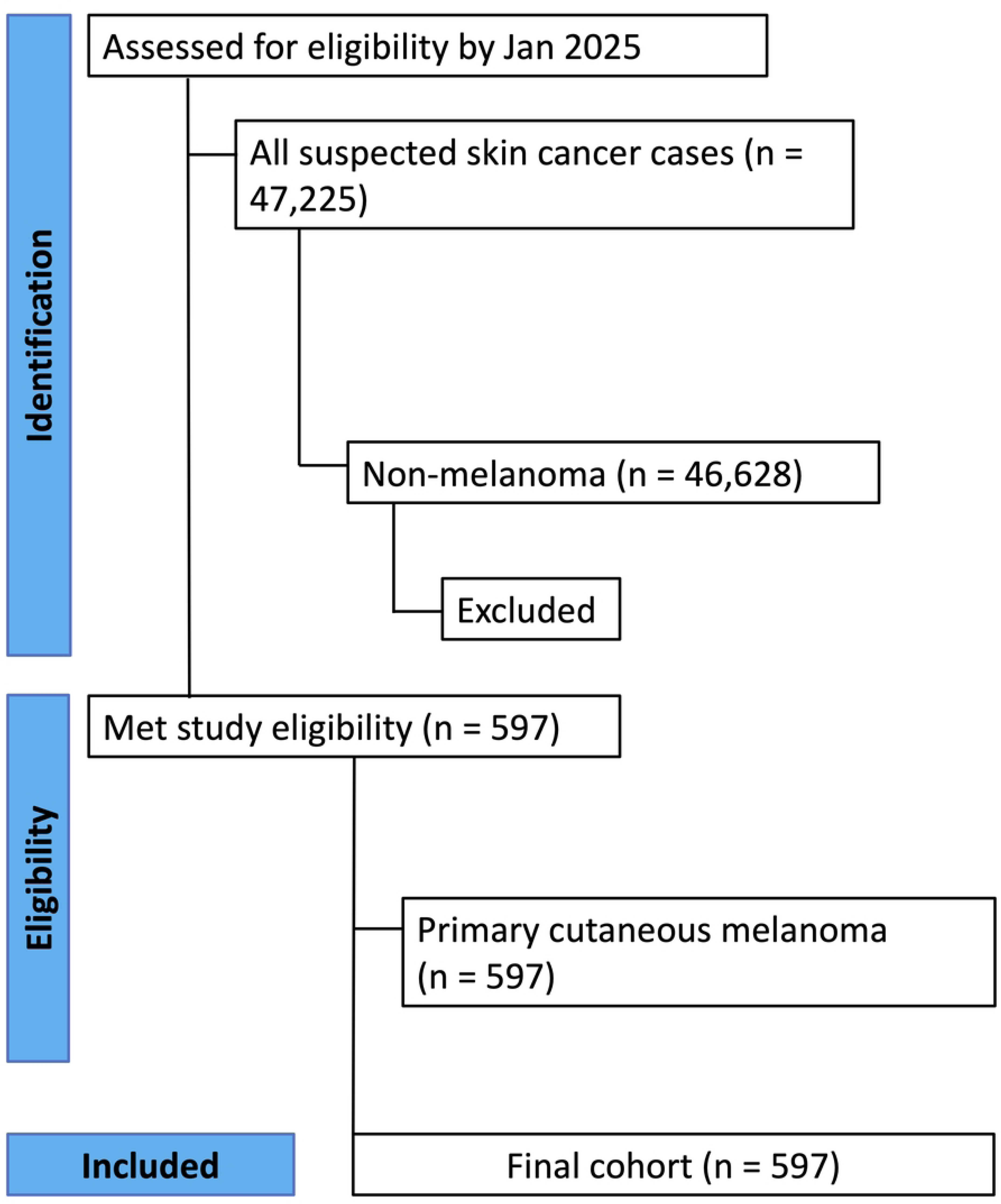
STROBE Flow diagram demonstrating identification, screening, and inclusion of patients with primary cutaneous melanoma.

**Table 1.** Baseline characteristics of the study population.

|  | **Total**<br>**(n = 597)** | **Males**<br>**(n = 303)** | **Females**<br>**(n =294 )** |
| --- | --- | --- | --- |
| **Age (years)** | 47.4 ± 12.3 | 50.63 ± 11.90 | 44.05 ± 11.97 |
| **Ethnicity** |  |  |  |
| European | 406 | 212 | 194 |
| MENA | 65 | 32 | 33 |
| African | 33 | 12 | 21 |
| Australasian | 28 | 21 | 7 |
| S/SE Asian | 11 | 3 | 8 |
| S American | 10 | 5 | 5 |
| Other | 44 | 18 | 26 |
| **Clark Level** |  |  |  |
| I | 121 | 58 | 63 |
| II | 221 | 104 | 117 |
| III | 172 | 93 | 79 |
| IV | 82 | 47 | 35 |
| V | 1 | 1 | 0 |

**Abbreviations:** MENA: Middle East and North Africa, S: South, SE: South East

### 3.2 Demographic Profile

The mean age at diagnosis for the entire cohort was 47.4 ± 12.3 years. A statistically significant difference in age at diagnosis was observed between the sexes, with males presenting at a mean age of 50.6 ± 11.9 years, approximately 6.5 years older than females, who had a mean age of 44.1 ± 12.0 years (*p <* 0.001), possibly suggesting delayed presentation due to lower dermatological awareness and/or different sun exposure safety habits amongst them.

The cohort was ethnically diverse, though predominantly composed of individuals of European descent (n = 406, 73.4%). The next largest group was patients from the MENA region (n = 65, 11.8%), followed by smaller representations from African (n = 33, 6.0%), Australasian (n = 28, 5.1%), South and Southeast Asian (n = 11, 2.0%), South American (n = 10, 1.8%), and others (n = 44, 7.4%) populations.

### 3.3 Outcome data

#### Histological Subtypes

The most prevalent histological subtype was SSM, accounting for 58.5% of all cases. This was followed by melanoma in situ (14.7%), LMM (13.2%), and NM (9.7%). While SSM was the most common subtype in both sexes, NM was notably more frequent in males (12.2%) compared to females (7.1%) **(Figure 2)**.

**Figure 2.**
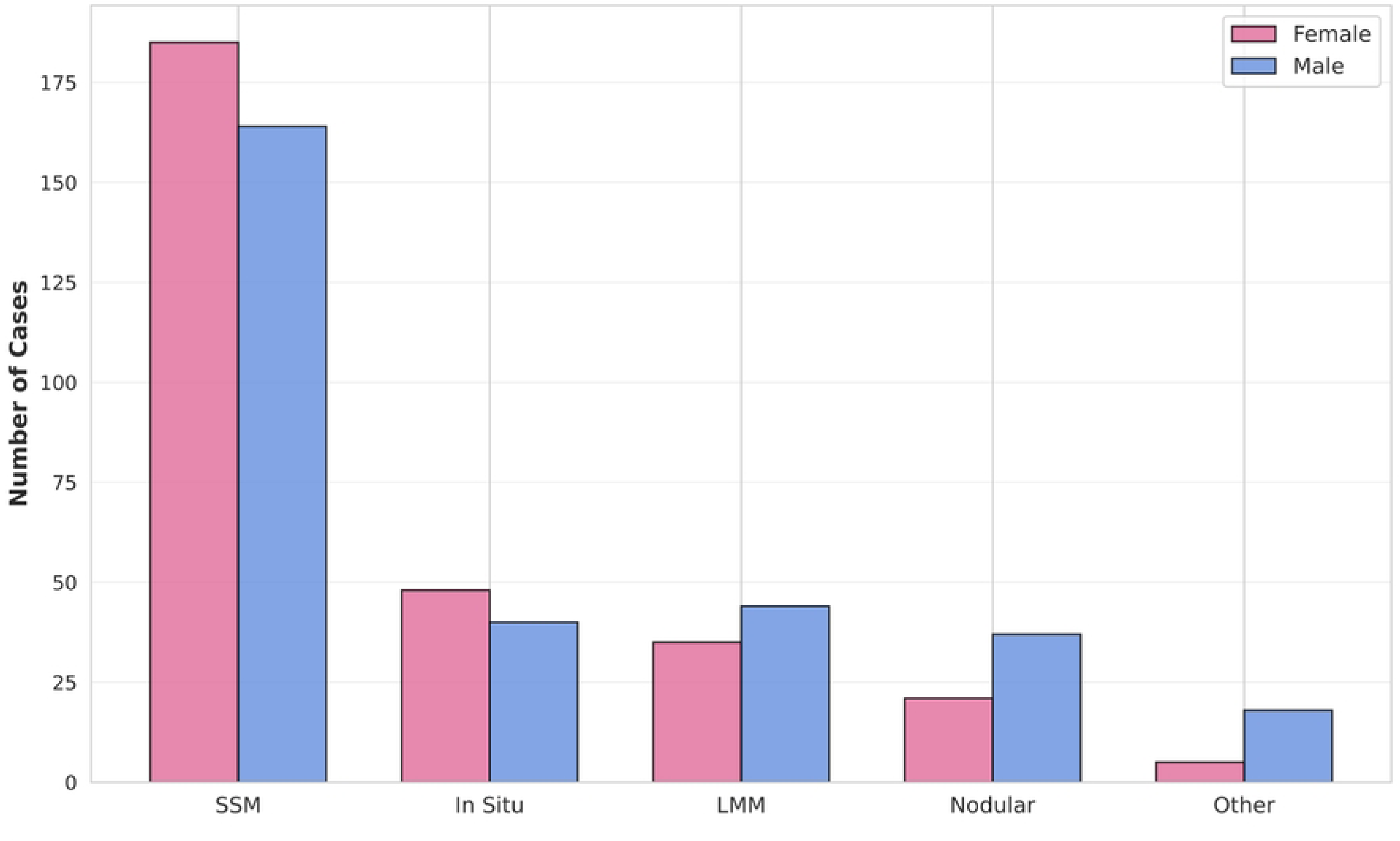
Distribution of Melanoma Histologic Subtypes by Sex. Other subtypes include acral lentiginous melanoma, desmoplastic melanoma, and spitzoid melanoma.

#### Tumor Invasion and Thickness

The distribution of the Clark invasion level is shown in **Table 1**. Most tumors were classified as Clark level II (37.0%) or III (28.9%). Regarding Breslow thickness, nearly half of the tumors (46.9%) were classified as thin (≤1.0mm), with an average thickness of 0.62 ± 1.03 mm. However, males had significantly thicker tumors on average than females (Breslow depth: 0.72 ± 1.32 mm vs. 0.50 ± 0.58 mm; *p <* 0.01). This trend is further reflected in the categorical distribution, in which a higher proportion of males presented with tumors >1.0 mm in thickness **(Figure 3)**.

**Figure 3.**
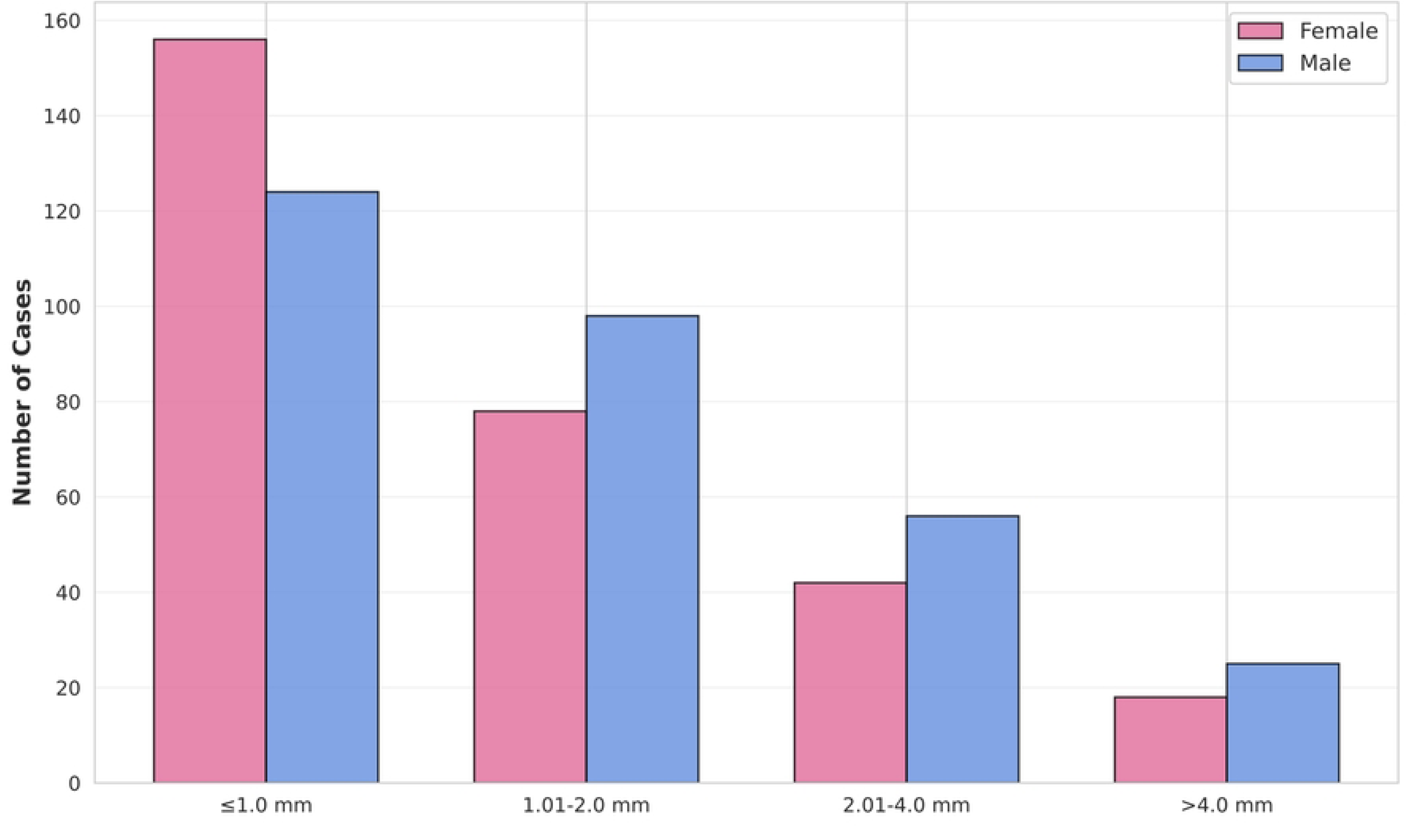
Breslow Thickness Categories of Melanoma by Sex.

#### Anatomical Location

The anatomical distribution of melanoma showed distinct patterns between sexes, as detailed in **Figure 4**. The most common location for melanoma in females was the leg and foot (37.8%), whereas in males, it was the back and torso (43.9%). Males also had a higher proportion of melanomas on the head and neck (15.8% vs. 8.8%).

**Figure 4.**
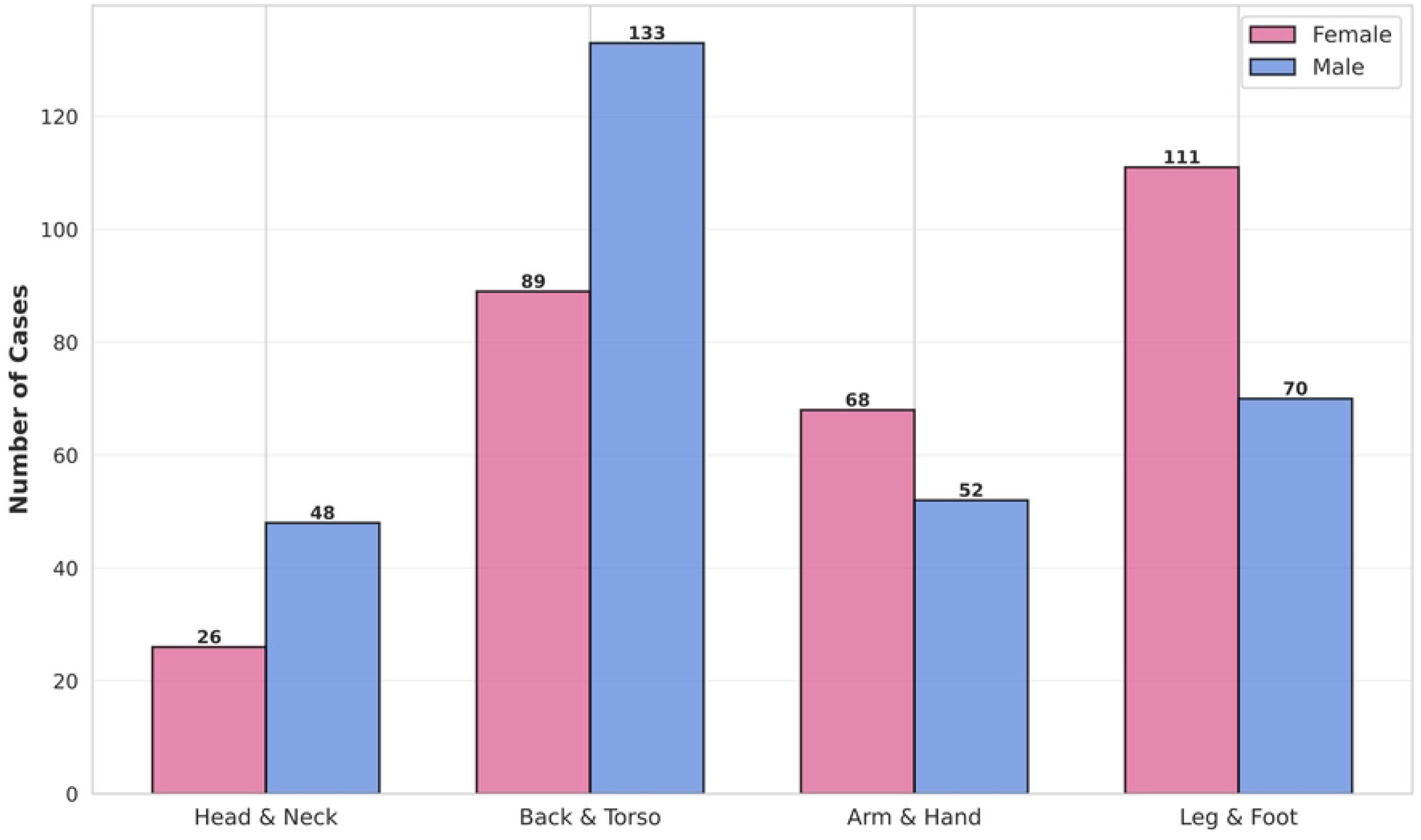
Anatomical Distribution of Melanoma Cases by Sex.

### 3.5 Subgroup Analysis – MENA Population

Among the 65 MENA patients identified in the cohort, melanoma presentation demonstrated distinct epidemiological and clinicopathological characteristics. Lebanese nationals accounted for the largest proportion of cases (40%), followed by Egyptians (16.9%), Emiratis (10.8%), Jordanians (7.7%), and Syrians (7.7%). The remaining nationalities (Palestinians, Bahrainis, Moroccans, Turks, and Omanis) contributed fewer than 5 cases. Sex distribution within the MENA subgroup showed a similar predominance (females 49.2%; males 50.8%), consistent with the overall sex distribution.

Histologic subtype analysis in the MENA subgroup demonstrated a predominance of SSM (58.5%), consistent with global patterns. The proportion of in situ melanomas in this subgroup was 16.9%, which is slightly higher compared to the overall cohort of 14.7% **(Figure 5)**.

**Figure 5.**
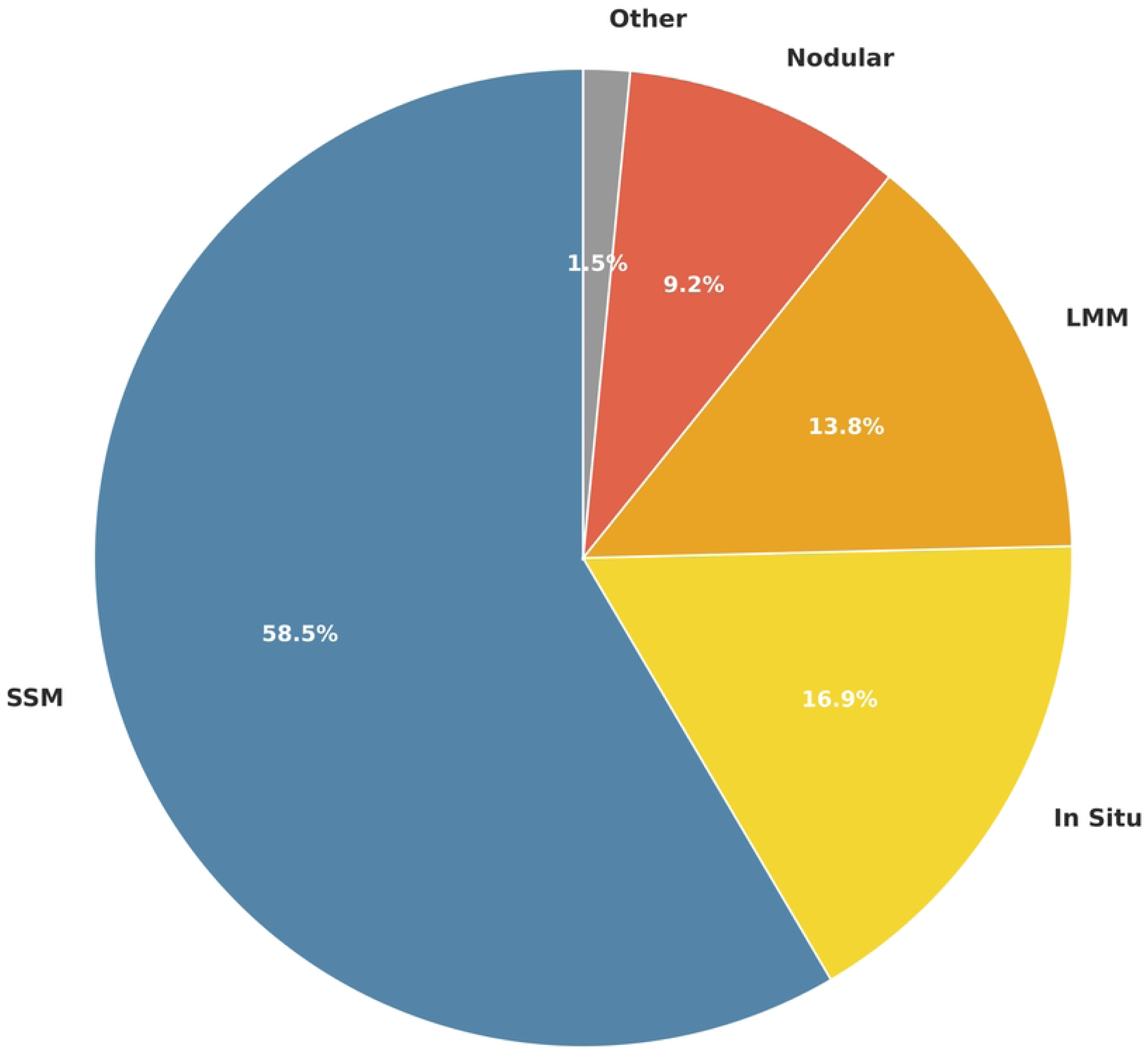
Histologic Subtype Distribution of Melanoma in the Middle East and North African (MENA) Subgroup.

Breslow thickness distribution in this subgroup of patients revealed that 40% presented with thin melanomas (≤1.0 mm), indicating a favorable prognosis. An additional 30.8% had intermediate thickness (1.01–2.0 mm), whereas 20% and 9.2% had thicker tumors (2.01–4.0 mm and >4.0 mm). The mean thickness in this subgroup was 1.11 ± 2.42 mm, which was comparable to the overall cohort mean of 0.62±1.03 mm (Mean Difference 0.49 mm; 95% CI [–0.10, 1.09]; *p =* 0.10)

### 3.4 Predictors of Thick Melanoma

To identify independent factors associated with a higher risk of presenting with a thick melanoma (Breslow depth >1.0 mm), a multivariable logistic regression analysis was performed. The results, presented in **Table 2**, identify two powerful, statistically significant predictors. Nodular histology was the strongest predictor, increasing the odds of a thick melanoma by over 18-fold compared to LMM (OR 18.40; 95% CI [7.08, 47.86]; *p <* 0.001). Similarly, each unit increase in Clark level of invasion was associated with a similar increase in the odds of a thick tumor (OR 18.50; 95% CI [8.44, 40.58]; *p <* 0.001). Other variables, including male sex, age, and anatomical site, were not found to be statistically significant independent predictors in this model.

**Table 2.** Multivariable logistic regression of predictors of thick melanoma (Breslow thickness >1.0 mm). Nodular melanoma subtype and Clark level were the only significant independent predictors.

| Variable | OR | 95% CI | P value |
| --- | --- | --- | --- |
| **Melanoma Type (Ref: LMM)** |  |  |  |
| Nodular | 18.402 | 7.705–47.859 | <0.001 |
| Superficial Spreading | 1.77 | 0.264–11.931 | 0.557 |
| Others | 2.145 | 0.518–8.879 | 0.295 |
| **Clark Level** | 18.503 | 8.438–40.557 | <0.001 |
| **Sex (Ref: females)** |  |  |  |
| Male | 1.465 | 0.582–3.689 | 0.418 |
| **Age group (Ref: <30)** |  |  |  |
| 30-39 | 0.259 | 0.043–1.558 | 0.14 |
| 40-49 | 0.958 | 0.177–5.192 | 0.961 |
| 50-59 | 0.447 | 0.076–2.642 | 0.374 |
| ≥60 | 0.958 | 0.165–5.559 | 0.962 |
| **Anatomical site (Ref: Leg & Foot)** |  |  |  |
| Head and Neck | 1.537 | 0.357–6.623 | 0.564 |
| Back and Torso | 0.706 | 0.240–2.075 | 0.526 |
| Arm and Hand | 0.624 | 0.198–1.967 | 0.421 |

**Abbreviations:** Ref: Reference, OR: Odds Ratio, CI: Confidence Interval, LMM: Lentigo Maligna Melanoma.

## 4. Discussion

This eight-year, multi-ethnic cohort study provides a comprehensive epidemiological analysis of cutaneous melanoma in the UAE, revealing a significant burden of disease, particularly among the nation’s large expatriate population. Our findings, which show a predominance of cases among individuals of European descent and distinct anatomical distribution patterns between sexes, reflect established global trends in the literature[10–12]. However, the primary significance of these epidemiological data lies in the urgent public health imperative they expose: a high-risk environment coupled with critically low levels of public awareness and engagement in preventive behaviors.

### The Epidemiological Burden and Global Context

Melanoma has historically been considered rare in most African and Asian countries, including the Middle East, with incidence rates often below 1 per 100,000 person-years[2]. However, this trend is being challenged by emerging epidemiological data from the region. In the UAE, skin cancer now ranks as the fourth most common malignancy, accounting for 6.3% of all malignant cases[5]. Our study’s findings that 73.4% of melanoma cases occurred in individuals of European descent reflect the nation’s unique demographic composition and the heightened vulnerability of fair-skinned expatriates to melanoma in the high-UV environment of the Arabian Peninsula.

The epidemiological patterns observed in our cohort are consistent with established global trends regarding Breslow thickness, histological subtypes, and anatomical distribution. SSM was the most prevalent subtype, followed by melanoma in situ, LMM, and NM, consistent with the international literature[1,11]. The distribution of Breslow thickness, with nearly 50% classified as thin (≤1.0 mm), is favorable, suggesting some degree of early detection. However, this proportion remains below the 53.5% reported in countries with established public awareness programs[13]. The mean Breslow thickness of 0.62 mm in the overall cohort is comparable to that in developed nations; however, the significant difference between males and females warrants particular attention, as it reflects disparities in screening practices and disease detection patterns[10,14,15].

### Regional Context: A Comparison with the MENA Subgroup

Our study provides a unique opportunity to contextualize the presentation of melanoma within the MENA region. A comparative analysis of our findings with data from Jordan and Iran reveals both similarities and important distinctions in the clinicopathological features of melanoma among different MENA populations. In our cohort, the mean Breslow thickness in the MENA subgroup was 1.12 ± 2.42 mm, which is notably lower than the 3.87 ± 3.35 mm reported by Malakoutikhah *et al.*[16]. This suggests that patients in the UAE may present with thinner tumors than those in other regions, potentially reflecting greater access to specialized dermatological care and higher health literacy in the UAE’s metropolitan centers. However, the mean Breslow thickness in our MENA subgroup remains notably higher than the overall cohort’s average of 0.62 ± 1.03 mm, indicating a need for targeted awareness campaigns in this population.

Furthermore, the histological subtype distribution varies across the region. While SSM was the most common subtype in our overall cohort, ALM is the most prevalent subtype in Iran, accounting for 36.5% of cases in one study[16]. In contrast, ALM constituted only 3.7% of cases in our overall cohort and was not the predominant subtype in our MENA subgroup. These differences highlight the heterogeneity of melanoma presentation across the MENA region and underscore the importance of tailoring public health messaging to each country’s specific epidemiological pattern.

### Sex and Gender Disparities in Melanoma Presentation

The higher incidence of melanoma in males and their presentation with thicker tumors is a well-documented phenomenon globally[10,14,15]. In our cohort, males presented at a mean age of 50.6 years, approximately 6.5 years older than females, and demonstrated a higher proportion of NM, a subtype associated with poor prognosis. The anatomical distribution also reflected sex-specific patterns, with males showing a higher proportion of melanomas on the back and torso, and head and neck, whereas females predominantly presented with leg and foot lesions. These differences have been attributed to behavioral and occupational factors, with males being more likely to engage in outdoor activities without adequate sun protection and to work in occupations with prolonged sun exposure[15,17]. Furthermore, gender differences in healthcare-seeking behavior and attitudes toward preventive care contribute to later diagnosis in males[15].

### The Critical Public Health Awareness Crisis in the UAE

The core challenge in the UAE is a disconnect between the high ambient ultraviolet (UV) radiation and the public’s perception of skin cancer risk. Our study confirms that melanoma is not a rare malignancy in the region, a finding that is sharply contrasted by the population’s inadequate knowledge and screening practices. Jarab *et al.*[5] quantified this gap, revealing that despite skin cancer being the fourth most common malignancy in the UAE, the public exhibits moderate knowledge, unfavorable attitudes, and deficient screening practices. The most significant barriers identified were a fundamental lack of knowledge about skin cancer (74.1%), unawareness of the need for screening (72.2%), and failure to seek medical evaluation in the absence of symptoms (54.1%)[5]. These statistics highlight a significant public health issue where the most critical tool for reducing melanoma mortality, early detection, is severely underutilized.

### Cultural, Environmental, and Healthcare System Barriers

The unique multicultural composition of the UAE, with its mix of sun-seeking expatriates and a local population that may have a false sense of security due to darker skin phenotypes, further complicates public health messaging and necessitates culturally appropriate, targeted interventions.

The traditional full-body coverage in Emirati culture (traditionally called the kandura for males and the abaya for females), while providing sun protection, may paradoxically reduce awareness of skin changes and the importance of regular skin self-examination[5]. Furthermore, a cultural preference for tanned skin achieved through unprotected sunbathing contributes to reduced awareness of UV-related risks, particularly in regions with intense sunlight[5]. Additionally, there is a widespread misconception that skin cancer primarily affects individuals with lighter skin tones, leading to an underestimation of risk among darker-skinned populations. However, ALM, which accounts for a larger proportion of melanomas in individuals with skin of color, is often diagnosed at a more advanced stage because it is often located on the palms, soles, and nail beds, where it is easily overlooked[18,19].

### Evidence for the Effectiveness of Public Health Interventions

Overcoming these barriers requires a strategic shift from passive information dissemination to active, evidence-based public health engagement. The success of such initiatives is well documented in the international literature[13,20]. In countries with established awareness programs, a significant increase in the diagnosis of thin melanomas and a corresponding reduction in mortality have been observed. A retrospective analysis by Armstrong *et al.*[13] of a 10-year public awareness campaign demonstrated a substantial increase in the proportion of thin melanomas diagnosed, from 35.4% in 2003 to 53.5% in 2012, indicating a clear shift toward earlier detection[13]. Similarly, Matsumoto *et al.*[20] documented that a primary care-based melanoma screening initiative was associated with increased detection of thin melanoma and melanoma in situ, raising awareness of the importance of systematic screening.

The evidence for the efficacy of educational interventions is compelling. Nadratowski *et al.*[21] conducted a randomized online study to assess the impact of different social media messages on melanoma awareness (melanoma warning signs and correctly identifying moles) and skin self-examination behavior. Participants viewed Facebook and Instagram-style posts targeting either knowledge about melanoma, self-efficacy for skin checks, both, or control content. The study revealed that the group exposed to knowledge messages scored significantly higher on mole detection than the group exposed to control/self-efficacy messages and warning sign knowledge. Self-efficacy messages increased confidence and intentions to perform skin checks compared with control/knowledge-only messages[21]. These findings suggest that targeted, evidence-based social media content can positively influence both melanoma knowledge and self-screening behavior, offering a promising, scalable approach to enhance public engagement in melanoma prevention and early detection efforts.

### Leveraging Digital Health and Social Media for Public Engagement

In the modern digital landscape, social media and mobile health technologies offer an unprecedented opportunity to deliver scalable and cost-effective educational interventions. The growing influence of these platforms is transforming health communication globally. The study by Nadratowski *et al.*[21] provides a template for developing effective digital interventions. The integration of artificial intelligence (AI) for dermoscopic analysis and skin lesion classification could serve as a cost-effective and time-saving educational tool, allowing individuals to assess suspicious lesions before seeking professional evaluation, thereby advancing the frontiers of melanoma screening[22,23].

School-based interventions represent another clinical avenue for prevention. Guy *et al.*[24] demonstrated that primary and middle school-based programs can significantly improve sun-protection knowledge and behaviors, with effects persisting into adulthood. These programs should include supportive policies allowing sunscreen use, environmental modifications to increase shade availability, and comprehensive education on skin cancer risk and prevention. Such interventions are critical in the UAE, where a large proportion of the population is young and where early establishment of protective behaviors could have lifelong benefits.

### Recommendations for a Comprehensive Public Health Strategy

Based on the evidence presented, we recommend a multi-pronged public health approach for the UAE:

1. National Awareness Campaign: Establishing a comprehensive, nationally coordinated skin cancer awareness campaign utilizing television, radio, print, and digital media. The campaign should be culturally sensitive, multilingual, and evidence-based.
2. Digital Health Integration: Develop and deploy mobile health applications and social media content targeting melanoma awareness and self-examination. These should incorporate interactive elements, dermoscopic images, and AI-assisted lesion assessment tools.
3. Healthcare System Integration: Integrate skin cancer screening into primary care settings and occupational health programs. Provide training to primary care physicians and occupational health nurses on melanoma recognition and referral protocols.
4. Targeted Interventions for High-Risk Groups: Develop targeted interventions for older adults, males, smokers, and uninsured populations to address the specific barriers these groups face in accessing screening.
5. School-Based Prevention: Implement comprehensive skin cancer prevention programs in primary and secondary schools, emphasizing sun protection and early detection.
6. Equity and Access: Ensure equitable access to dermatological expertise and screening services across all socioeconomic strata and immigrant populations, addressing the social determinants of health that influence melanoma outcomes.
7. Data Collection and Registry: Establish a national melanoma registry to track incidence, outcomes, and the impact of public health interventions, enabling evidence-based refinement strategies over time.

### Limitations

This study poses several limitations. Its reliance on data from a single private tertiary referral center may not capture the full spectrum of melanoma cases across the public health care sector, potentially limiting the generalizability of our findings. The underrepresentation of certain MENA nationalities and potential underreporting among migrant populations with limited healthcare access are also acknowledged. The study’s design precludes assessment of temporal trends or causal relationships. Future research should aim to integrate data from the public healthcare sector to create a more complete national registry. Longitudinal studies are needed to track incidence trends, treatment outcomes, and the impact of future public health interventions. Furthermore, incorporating molecular profiling of tumors could identify genetic factors prevalent in the region’s diverse population, paving the way for more personalized risk stratification and treatment.

## 5. Conclusions

This study highlights the epidemiological landscape of melanoma in the UAE and, more importantly, frames it as a pressing public health issue demanding immediate action. The confluence of a high-risk solar environment and a population with significant knowledge and awareness deficits creates a dangerous gap between risk and prevention action. The UAE has an opportunity to become a regional leader by implementing a comprehensive, multi-pronged public health strategy that combines robust national awareness campaigns, innovative digital health interventions, targeted educational programs to enhance health literacy, and equitable access to screening services. By empowering its diverse population with the knowledge and tools for early detection, the UAE can significantly reduce melanoma-associated morbidity and mortality and set a new standard for skin cancer prevention in the Middle East.

## Statements and Declarations

### Institutional Review Board Statement

This study was conducted in accordance with the Declaration of Helsinki, and the protocol was approved by the Dubai Scientific Research Ethics Committee (Reference number: DSREC-11/2024_41).

### Data availability statement

The datasets generated and analyzed during the current study are available from the corresponding author on reasonable request.

